# The Global Status of Global Surgery Indicators: A Scoping Review Protocol

**DOI:** 10.1101/2025.02.10.25321873

**Authors:** Gabriella Y. Hyman, Lauren Kratky, Ramya Reddy, Paul A. Bain, Nakul P. Raykar

## Abstract

**Background:** Surgical care is essential to achieving health system goals, including Universal Health Coverage (UHC) and the Sustainable Development Goals (SDGs). The Lancet Commission on Global Surgery (LCoGS) proposed core indicators to assess surgical system capacity, delivery, and financial risk protection. Despite their importance, variability and inconsistent reporting limit their utility in tracking progress toward Global Surgery 2030.

**Aims:** This review aims to quantify the status of five global surgery indicators globally, describe current indicator reporting practices, and identify facilitators and barriers to regular indicator reporting, providing recommendations to improve future tracking and policy implementation.

**Methods:** Following the Joanna Briggs Institute (JBI) and PRISMA-ScR guidelines, we will search peer-reviewed databases and grey literature to identify studies reporting LCoGS indicators. Data extraction will focus on indicator status, reporting frequency, and associated challenges. Results will be analyzed quantitatively and thematically.

**Conclusion:** By mapping current indicator reporting, this review will provide actionable insights to inform future policy, improve data collection practices, and accelerate progress toward achieving Global Surgery 2030 targets.

## Introduction

Surgical care is an essential component of any functioning health system (1–3). Conditions requiring surgical care account for nearly one-third of the global disease burden, meaning health equity is not achievable without access to safe, timely, affordable surgical care (3,4). These conditions include non-communicable diseases such as cancers, trauma and injury, and maternal and child health (4). Effective surgical systems are essential for reducing preventable morbidity and mortality, and achieving broader health system goals of universal health coverage (UHC) (3,5). Programs and policy interventions for strengthening surgical systems, such as National Surgical Plans (NSPs), rely on robust monitoring and evaluation frameworks to track progress and achieve targets. Indicators serve as measurable benchmarks to assess health system performance and tracking progress towards policy targets (6,7). Data collected from indicator tracking can be used as evidence to advocate for scarce human and capital resources needed to strengthen health systems and achieve the health-related Sustainable Development Goals (SDGs), which also converge in 2030 (8,9). Unfortunately, the adoption and reporting of global surgery indicators have been inconsistent across countries, despite their anticipated value (3,7,10).

In 2015, the Lancet Commission on Global Surgery (LCoGS) published a formative analysis on the global status of access to surgical care with corresponding targets for achieving *Global Surgery 2030* (3). The commission identified six core indicators essential for achieving equitable surgical care access. The indicators span the domains of health service delivery, workforce, information systems, and economic impact. The LCoGS indicators reflect health system readiness and capacity to deliver high-quality surgical care to all when needed. Through an extensive consensus development process, these indicators were selected for their alignment with the core elements of UHC (access to surgical care, the provision of essential surgical services, and financial risk protection). The indicators provide insight into the preparedness of the health system to provide surgical services (geographic access to care, surgical workforce density), service delivery (surgical case volume provided, perioperative mortality rate), and the impact of surgical care on patients (catastrophic health expenditure) (10). Additionally, the indicators align with the WHO Health System Building Blocks framework (11). Many of these indicators have been incorporated into the World Bank’s World Development Indicators (WDI) and the WHO 100 Core Global Health Indicators (6,12).

While the LCoGS demonstrated the importance of the core surgical indicators for showing the capacity of a health system to provide surgical services, evidence is needed to determine if improvements in surgical care capacity, delivery, and quality/impact on population health have been made. The absence of high-quality, country-level indicator data makes it almost impossible to meaningfully track progress towards global surgery targets and evaluate whether the targets laid out 10 years ago were appropriate and achievable at the outset. We are also unable to validate the current targets as they may over- or underestimate surgical need, capacity and quality through the generalized but not necessarily generalizable approach. Thus, there are two major gaps in our current knowledge of the global surgery indicators: we need an updated quantification of the current status of each indicator, and we need to improve our understanding of the practices surrounding data collection and reporting by all WHO member states. This is crucial for understanding the current state of global surgery and targeting interventions to improve surgical care access and meet the 2030 targets (Figure 1). This article outlines a protocol for a scoping review which aims to collect data regarding indicators and reporting, thus benchmarking the progress that has been made since the LCoGS in 2015. This can be used as a call to action to drive progress toward achieving the goal set for Global Surgery 2030.

**Figure 1.**
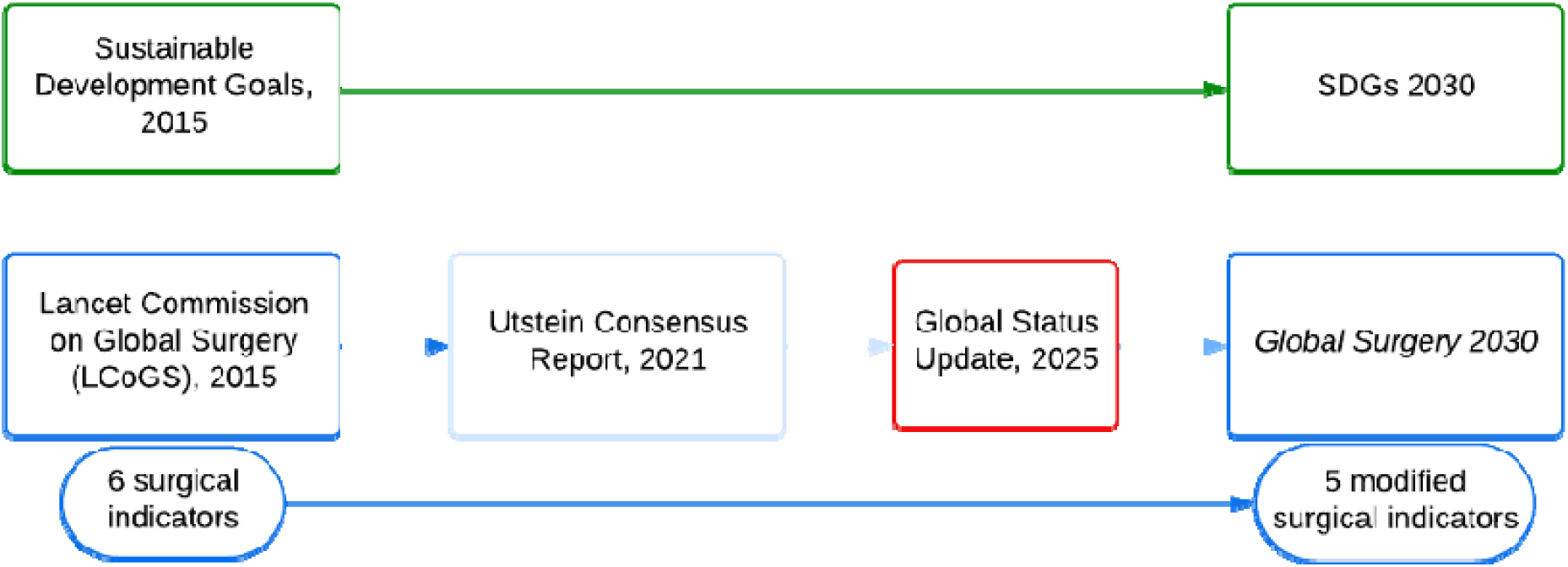
Timeline Towards Global Surgery 2030 Targets

In response to challenges in indicator reporting and tracking in the first 5 years following the LCoGS, the global surgery community, with both HIC and LMIC stakeholders, undertook an iterative process to revisit and refine the indicators. The indicator definitions and proposed methods for collection were updated in an Utstein process for developing and reporting guidelines (Table 1) (10). The major changes included condensing the two economic impact indicators of catastrophic and impoverishing financial consequences of surgery into one indicator (catastrophic health expenditure) and recommending changes to the access to timely essential surgery indicator to a geospatial access model that determines timeliness of access to a facility that has capacity to deliver surgery and anesthesia care for Bellwether procedures. The target for POMR remains the reporting rate itself (100% reporting by 2030) as opposed to a safety threshold. This likely reflects the paucity of data linked to case volumes and outcomes.

**Table 1:** Surgical indicator definitions, as of 2025 (3,10)

| *Indicator* | **LCoGS**<br>**Definition** | **LCoGS**<br>**Target** | **Utstein**<br>**Revised**<br>**Definition** | **Utstein**<br>**Target** | **Numerator** | **Denominator** | **Methods** |
| --- | --- | --- | --- | --- | --- | --- | --- |
| *Access* | Proportion of the population that can access, within 2 hours, a facility that can perform Bellwether procedures. | 80% access by 2030. | Proportion of the population within 2 hours of a facility capable of essential surgery, determined through geospatial analysis. | 80% access by 2030. | Number of people within 2 hours of a capable facility. | Total population of the country. | Various methods for calculating access, including Geographic Information System (GIS), drive-time models, and population data. Consider stratification by surgery type, equity, and facility capacity. |
| *Surgical Workforce Density* | Number of surgical, anaesthetic, and obstetric specialists working per 100,000 population. | 20 SAO providers per 100,000 population by 2030. | Number of active surgical, anaesthetic, and obstetric providers per 100,000, including certified non-physician providers. | 20 SAO providers per 100,000 population by 2030. | Number of SAO providers actively working in the country. | Population size (per 100,000) | Should differentiate between specialists and non-specialist providers. |
| *Surgical Volume* | Number of surgical procedures performed in operating theatres per 100,000 population per year. | 5,000 procedures per 100,000 population per year by 2030. | Number of procedures performed in operating theatres per 100,000 population per year, with standardized definitions. | 5,000 procedures per 100,000 population per year by 2030. | Total number of surgical procedures performed annually. | Population size (per 100,000). | Practices vary widely in terms of who reports and the granularity of reporting (eg by procedure type). |
| ***Perioperative Mortality Rate (POMR)*** | All-cause mortality before discharge among patients who have undergone a procedure in an operating room, expressed as a percentage of total procedures per year | 100% tracking by 2030. | All-cause mortality before discharge (before 30 days) among patients who received anaesthesia for a procedure in an operating room, divided by the total number of procedures, per year, expressed as a percentage. | 100% tracking by 2030. | Number of post-operative, in-hospital deaths, capped at 30 days. | Total number of procedures performed. | Common variations in reporting include differences in follow-up periods and definitions of mortality. |
| ***Protection Against Catastrophic Expenditure*** | Proportion of households protected against catastrophic expenditure from out-of-pocket payments for surgical and anaesthesia care. | 100% protection by 2030 | Percentage of households protected against catastrophic expenditure (threshold of 10% of total household expenditure) from direct out-of-pocket payments for surgical care | 100% protection by 2030 | Number of households not incurring catastrophic health expenditure | Total number of households | Requires household-level data; often modeled due to lack of primary data |
| ***Protection Against Impoverishing Expenditure*** | Proportion of households protected against impoverishment from out-of-pocket payments for surgical and anaesthesia care | 100% protection by 2030. | Combined with catastrophic expenditure to measure catastrophic health expenditure |  |  |  |  |

The impact of the 2021 indicator definition updates on data collection, data reporting, and surgical policy is unclear. For example, a systematic review of global surgery indicators within India, published in June 2023, showed that India did not meet the targets set by LCoGS for any of the indicators and that there was limited data for certain indicators-specifically geospatial access and economic impact (13). Additionally, there was variability in the evidence available for workforce density, surgical volumes, and POMR. This study showed a need for subnational district and state level data reporting as well as reporting between the public and private health sectors and different levels of care (13). Cases of successful indicator reporting appear to be linked to structured national or regional initiatives. For example, Brazil’s Unified Health System, (Sistema Único de Saúde, SUS), has implemented a robust data collection system that enables consistent reporting of surgical indicators (14). Other examples are linked closely to efforts to conduct a baseline assessment for the NSP development. Such was the case in the Pacific Region where 14 countries collected data on the first four of six indicators with data collection support from the Royal Australasian College of Surgeons and the Pacific Islands Surgical Association (15). In Ecuador and Namibia, similar nationwide baseline assessments that generated the indicators were conducted for the purpose of developing an NSP (16). Other countries appear to rely on retrospective record reviews to collect indicator data. For example, in Uganda, a standardized quantitative assessment and semi structured interview was administered to key stakeholders at 17 randomly selected public hospitals in order to gain insight on surgical indicator data (17). Interestingly, indicator reporting in HICs does not appear much better. The data relevant to the indicators may be accessible via ministries of health, other professional societies, or in reviews conducted by individuals groups but this data is not often synthesized and reported at a national level (12).

The variability and sporadic nature of surgical indicator reporting within and across countries shows a need to improve our understanding about current practices (7). The recommendation in LCoGS was for annual reporting to enable consistent monitoring and evaluation of NSPs and health system strengthening in general. Challenges in reporting also stem from the heterogeneity in the indicator definitions and methodologies (Table 1). We expect many factors cited as facilitators and barriers to NSP development, such as financing, stakeholder engagement and political will, affect surgical indicator reporting as well (18). While much of the existing data mentioned is reported at a subnational level, or not routinely reported, understanding the methods that have facilitated success may help us scale up and expand efforts to understand the status of surgical indicators and access.

Given the need for evidence-based policies, data-driven health systems strengthening, and the fast-approaching Global Surgery 2030 targets, the scoping review aims to provide a comprehensive status update on progress towards equitable surgical care access by quantifying the core indicators. The review will provide critical insights into reporting practices, including where reporting takes place, how and where reporting occurs, how the indicators are used, and facilitators and barriers to reporting.

Additionally, the review aims to provide recommendations to inform future policy development, increase reporting rates and help develop strategies for data collection and surgical care access (7). Improved indicator reporting has the potential to accelerate our progress towards Global Surgery 2030 goals and to establish more effective and equitable targets for the future.

### Study aim

The overall aim of this scoping review is to map the available literature that pertains to LCoGS indicator reporting and provide a status update on each indicator globally. The secondary objectives are to describe practices surrounding indicator reporting and synthesize recommendations.

## Methods

We will use the methods outlined in the Joanna Briggs Institute (JBI) manual for knowledge synthesis and the Preferred Reporting Items for Systematic Reviews and Meta-Analysis extension for Scoping Reviews (PRISMA-ScR) guidelines to use literature across study designs in both peer-reviewed (indexed) databases and grey literature (19,20). The study will be completed in five phases: (1) determining the search question; (2) search strategy, (3) inclusion criteria, (4) data extraction, and (5) analysis and presentation of the results. The study will take place from 2024 to 2025. Institutional Review Board (IRB) approval is not required to conduct this study.

### Determining the research question

The research team convened a group of experts with experience developing and evaluating the LCoGS indicators. Many in the group were involved in the initial systematic reviews, consensus development, and synthesis studies when the indicators were first proposed in 2015 and reviewed in 2021. During the meetings, the group raised questions about the current status of surgical indicators and discussed the various proposed and current methods for determining each indicator. The group reflected on the process surrounding the first ‘status report’ for each indicator at the time of LCoGS and reflected on the documented and experienced challenges in reporting and monitoring the indicators. The group focused on possible facilitators and barriers to collecting and reporting indicator data and reflected on experiences from different regions where members have directly been involved in indicator collection, review or analysis. Additionally, the group considered the value of core indicator reporting for global surgery stakeholders and questioned whether an assessment of the appropriateness of the targets, and progress towards the targets, can be made without first conducting a comprehensive status update. Based on the feedback sessions, the review aims to answer the following question “*What is the status of access to safe, timely, affordable surgical care globally, according to global surgery indicators*?”

The objectives are:

- *To quantify each global surgery indicator: access, case volume, workforce density, perioperative mortality rate, and catastrophic health expenditure*.
- *To determine and describe current practices surrounding LCoGS indicator reporting*
- *To determine the feasibility of routine reporting*
- *To identify successful practices in indicator reporting to synthesize recommendations for future LCoGS indicator reporting*

### Search Strategy

Relevant peer-reviewed studies will be identified by searching the electronic databases MEDLINE (Ovid), Embase (Elsevier), Web of Science Core Collection (Clarivate), Global Health (EBSCO), and the Global Index Medicus (WHO). An independent search for each of the indicators has been developed by a medical librarian (PAB). Each search includes terms for the indicator and terms for surgery or surgical specialties (Appendix 1 includes MEDLINE searches for each indicator). Appropriate controlled vocabulary terms are included when available. For some of the indicators (surgical volume, workforce, and mortality rate), the searches are limited by the denominators used to report population level measures (*e*.*g*., 100,000; 10,000) to reduce retrieval to a manageable level. The searches also capture any study record that mentions Lancet indicators specifically. No date or language limits are applied to the searches.

A grey literature search will be conducted to identify non-indexed literature of relevance. Data sources, including but not limited to databases, registries and dashboards will be used to identify indicators for all countries. The studies included in the review will be managed and screened in Covidence (Covidence systematic review software, Veritas Health Innovation, Melbourne, Australia. Available at www.covidence.org.). The results set for each indicator will be screened independently with independent inclusion criteria for each indicator.

### Inclusion Criteria

The inclusion criteria for this scoping review uses the JBI definitions for population, concept, and context (20).

#### Population

Our review focuses on all populations. Any literature that reports at least one of the 5 LCoGS indicators for populations of all ages, sex, ethnicity or other demographic variables will be considered for inclusion. As the focus of the review is primarily on quantifying the indicators, the review will consider publications that report a critical component of the indicator, such as the numerator. Since we anticipate a low number of nationally representative publications, we will include publications that report data at a national, sub-national or regional, and single facility level for manual estimation of the indicator where possible.

#### Concept

The main concept for our review is the quantification of the LCoGS indicators of access to safe, timely, affordable surgical care. The LCoGS indicators were first defined in the LCoGS in 2015. We will use the most recent, modified definitions developed through an Utstein process in 2021 as these are the most widely-accepted by the policy and academic global surgery communities (Table 1). To ensure consistency studies will be included in their descriptions mentioning either the LCoGS, the Utstein indicators, or both.

The secondary concept pertains to the practices surrounding indicator reporting. This is described as possible facilitators or barriers to reporting global surgery indicators. If a study’s focus is on practices surrounding indicator reporting, the study will only be considered for inclusion if the study presents data that quantifies an indicator.

#### Context

This review will consider global surgery indicator reporting across geographic and health system settings. Any publication reporting at least one LCoGS indicator, in at least one WHO Member state will be eligible for inclusion. In addition to producing a global status report, these indicators serve the foundation for reporting and monitoring progress towards WHA Resolution 68.15 and more recently WHA 72.15 (21,22).

### Date range

All relevant publications published before the end of 2024 will be included in the review.

### Types of Sources

This scoping review will consider a broad range of literature sources reporting any quantitative, qualitative, mixed-method, experimental (e.g clinical trials) or observational (e.g. cohort or cross-sectional) methods. We will include any study design. Qualitative studies will also be considered provided they present data relevant to both the primary and secondary objectives of this review. Grey literature will include unpublished research, conference abstracts, policy documents, databases, organizational reports and proceedings. Literature will be limited to publications in English.

### Data management

Once all identified sources have been extracted from all databases, duplicates will be removed. Data will be uploaded and stored in Covidence for screening. The inclusion and exclusion criteria will be tested randomly for each indicator. Regarding abstract and title screening for each indicator, at least one of 2 experts for a particular indicator, assigned *a priori*, will be required to screen every source. Conflicts will be resolved by a third member of the core research team. For full text screening, at least one of 2 experts for a particular indicator, assigned *a priori*, will be required to screen every source. Reasons for exclusion will need to be given in Covidence. These will include: no English full text, abstract only, full text not available, duplicate, does not measure indicator. REDCap will be used for data extraction.

### Data extraction

A copy of each source article or document will be obtained, reviewed, and charted by two research team members. Specified team members for each indicator will extract data using a data extraction form that will be piloted before. The data extraction sheet may be modified following the pilot and any modifications will be detailed in the full scoping review report. Table 2 outlines the data extraction domains for the study.

**Table 2:** Data extraction domains.

| Domain | Variables to be extracted |
| --- | --- |
| Publication information | Author/ stakeholder, author affiliation, publication type, publication year/s, location of research (eg: country, World Bank income classification), data source (eg: journal, database, website, report, policy etc) |
| Scope of publication | Generalizability (international, national, subnational, multicenter, single center), number of facilities, publication timeline |
| Indicator components | Numerator, denominator, population size, source of population data, indicator format (eg: count, proportion, ratio, mean), calculation required, number of indicators reported (eg: to calculate trends; level of reporting by geography), relevant covariates reported (eg: to allow for stratification and modeling) |
| Methodology | Publication design, primary or secondary data source, method used for calculating indicator, date/ year of indicator |
| Reporting practices | Frequency of reporting (eg: once-off, annually), stakeholders conducting reporting |
| Feasibility | Funding (eg: source, amount, frequency), policy alignment, technical capacity (eg: electronic medical record, vital statistics, national dataset, dedicated team) |

### Critical appraisal of literature

As the primary aim of this scoping review is to provide an update on surgical indicators and reporting practices using existing literature, we will not undertake a formal qualitative assessment of the quality of each included literature source. However, we will document the study design, methods used for indicator data collection, calculation and reporting, and the method for manual calculation if the study team undertook additional analysis (for example for count data).

### Analysis

We will present our results in a modified PRISMA-ScR diagram, provide a basic numerical count for the type and characteristics of included literature sources, and conduct a quantitative analysis and visualization of the extracted data (23). We will produce a geographic map of the indicators and where possible, describe trends in indicator values. Additionally, we will analyze extracted data on literature authorship, funding, methodologies and publication types to answer the question of reporting facilitators and barriers.

## Data Availability

All data produced in the present work are contained in the manuscript

## Data Validation

We anticipate that we will produce a strategy and data extraction framework that can be used for regular reviews and global status updates of the current global surgery indicators.

## Appendix 1 MEDLINE (Ovid) searches for each LCoGS indicator

Case Volume

1. exp workload/ or (((surgical or surgery or surgeries or neurosurg* or operative) adj6 (caseload? or case-load? or volume? or workload?)) or ((surgical or surgery or neurosurgical or ophthalmol* or urolog* or gyn?ecol* or obstetric* or orthop?edic or operative) adj cases) or operative-rate? or “operations 100-000” or “procedures 100-000” or “operations-100000” or “procedures-100000” or ((surgeries or procedures) adj6 (“per-year” or annual* or annum or number or “12 month?” or “twelve-month?”))).ab,kf,kw,ti.
2. exp specialties, surgical/ or exp surgical procedures, operative/ or surgery.fs. or (surgery or surgeries or surgical or operative or an?sthesia or surgeon? or an?sthesiologist? or an?sthetist? or obstetric* or gyn?ecolog* or neurosurg* or ophthalmolog* or urolog* or orthop?edic*).ab,kf,kw,ti.
3. (“1000” or “10000” or “100000” or 1-000 or 10-000 or 100-000 or thousand).ab,kf,kw,ti.
4. ((lancet-commission adj3 global-surgery) or lcogs or lancet-indicator* or surgical-indicator?).ab,kf,kw,ti.
5. (1 and 2 and 3) or 4

Workforce

1. exp workforce/ or (((surgical or surgeries or operative) adj6 capacit*) or workforce or manpower or human-resource? or staff or staffing or labo?r-force).ab,kf,kw,ti.
2. exp specialties, surgical/ or exp surgical procedures, operative/ or surgery.fs. or (surgery or surgeries or surgical or operative or an?sthesia or surgeon? or an?sthesiologist? or an?sthetist? or obstetric* or gyn?ecolog* or neurosurg* or ophthalmolog* or urolog* or orthop?edic*).ab,kf,kw,ti.
3. (“1000” or “10000” or “100000” or 1-000 or 10-000 or 100-000 or thousand).ab,kf,kw,ti.
4. (provider-densit* or surgeon-densit* or specialist-densit* or provider-population).ab,kf,kw,ti.
5. ((lancet-commission adj3 global-surgery) or lcogs or lancet-indicator* or surgical-indicator?).ab,kf,kw,ti.
6. (1 and 2 and 3) or (4 and 2) or 5

Mortality Rate

1. (exp postoperative period/ and exp mortality/) or ((operative or perioperative or postoperative or surgery or surgical) adj3 (mortalit* or death?)).ti,kf,kw,ab.
2. exp specialties, surgical/ or exp surgical procedures, operative/ or surgery.fs. or (surgery or surgeries or surgical or operative or an?sthesia or surgeon? or an?sthesiologist? or an?sthetist? or obstetric* or gyn?ecolog* or neurosurg* or ophthalmolog* or urolog* or orthop?edic*).ab,kf,kw,ti.
3. (“1000” or “10000” or “100000” or 1-000 or 10-000 or 100-000 or thousand).ab,kf,kw,ti.
4. pomr.ab,kf,kw,ti.
5. ((lancet-commission adj3 global-surgery) or lcogs or lancet-indicator* or surgical-indicator?).ab,kf,kw,ti.
6. (1 and 2 and 3) or (4 and 2) or 5

Catastrophic Expenditure

1. ((catastrophic or impoverish*) adj6 (expen* or payment? or cost or costs)).ab,kf,kw,ti.
2. exp specialties, surgical/ or exp surgical procedures, operative/ or surgery.fs. or (surgery or surgeries or surgical or operative or an?sthesia or surgeon? or an?sthesiologist? or an?sthetist? or obstetric* or gyn?ecolog* or neurosurg* or ophthalmolog* or urolog* or orthop?edic*).ab,kf,kw,ti.
3. ((lancet-commission adj3 global-surgery) or lcogs or lancet-indicator* or surgical-indicator?).ab,kf,kw,ti.
4. (1 and 2) or 3

Timely Access

1. ((timely adj3 access*) or ((2-hour or 2-hr or 2-hrs or two-hour or 120-minute? or 60-minute?) adj3 (threshold or distance or access or radius or travel-time? or surgical facilit?))).ab,kf,kw,ti.
2. exp specialties, surgical/ or exp surgical procedures, operative/ or surgery.fs. or (surgery or surgeries or surgical or operative or an?sthesia or surgeon? or an?sthesiologist? or an?sthetist? or obstetric* or gyn?ecolog* or neurosurg* or ophthalmolog* or urolog* or orthop?edic*).ab,kf,kw,ti.
3. ((lancet-commission adj3 global-surgery) or lcogs or lancet-indicator* or surgical-indicator?).ab,kf,kw,ti.
4. (1 and 2) or 3

